# COVID-19 data reporting systems in Africa reveal insights for pandemic preparedness

**DOI:** 10.1101/2021.10.01.21264385

**Authors:** Seth D. Judson, Judith Torimiro, David M. Pigott, Apollo Maima, Ahmed Mostafa, Ahmed Samy, Peter Rabinowitz, Kevin Njabo

**Affiliations:** Department of Medicine, University of Washington, Seattle, USA; Faculty of Medicine and Biomedical Sciences, University of Yaoundé I, Cameroon; Department of Health Metrics Sciences, University of Washington, Seattle, USA; School of Pharmacy, Maseno University, Kenya; Center of Scientific Excellence for Influenza Viruses, National Research Centre, Giza, Egypt; Reference Laboratory for Veterinary Quality Control on Poultry Production, Animal Health Research Institute, Agricultural Research Center, Giza, Egypt; Immunogenetics, The Pirbright Institute, Surrey, UK; Departments of Environmental and Occupational Health Sciences, Global Health, University of Washington, Seattle, USA; Center for Tropical Research, University of California, Los Angeles, USA

## Abstract

**Background:** Throughout the coronavirus disease 2019 (COVID-19) pandemic, there have been a variety of practices for international reporting of COVID-19 data. African countries have used different national reporting systems to publicly share data. Analyzing the content, format, and frequency of these systems could elucidate lessons for future pandemics.

**Methods:** We examined national COVID-19 reporting practices across 54 African countries through 2020. Reporting systems were compared by type of report, frequency, and data content. We also compared reporting of metrics such as a patient demographics and co-morbidities, healthcare capacity, and diagnostic testing. We further evaluated regional and country-specific reporting practices in Cameroon, Egypt, Kenya, Senegal, and South Africa as examples from different sub-regions.

**Results:** National COVID-19 reporting systems were identified in 53/54 (98.1%) countries in 2020. Reporting systems were diverse and could be categorized into social media postings, websites, press releases, situation reports, and online dashboards. Of countries with reporting systems, 36/53 (67.9%) had recurrent situation reports and/or online dashboards which provided the highest quality of data.

**Conclusions:** African countries created diverse reporting systems to share COVID-19 data. Many countries used routinely updated situation reports or online dashboards. However, few countries reported patient demographics, co-morbidities, diagnostic testing practices, and healthcare capacity. Including these metrics as well as improving standardization and accessibility of data reporting systems could augment research and decision-making, as well as increase public awareness and transparency for national governments.

## Background

The COVID-19 pandemic has revealed global differences in pandemic preparedness and response. A major challenge has been the variability in reporting of COVID-19 data among countries. Accurate and timely reporting of COVID-19 cases, healthcare capacity, and epidemiological risk factors are crucial for guiding national policies, international decision-making, predictive modeling, and risk assessment. Public reporting of national data is also important for government transparency and trust. Researchers and policymakers can also promptly analyze publicly available data in order to inform public health interventions. As we continue to confront the COVID-19 pandemic and prepare for future pandemics, it will be important to examine national data reporting practices to find areas for improvement.

Given the unprecedented global scale and rapid progression of the COVID-19 pandemic, countries have had to quickly develop methods for reporting and assessing COVID-19 data. Researchers and policymakers have faced challenges in analyzing these international data given variability and instability in reporting, which has led to a call for a standardized approach for data reporting (1). An analysis of COVID-19 data reporting within the United States showed large variability in data reported between states and highlighted the lack of transparent and standardized COVID-19 data (2). Internationally, a wide variety of reporting systems have been used by different countries, including a diverse variety of online dashboards which differ in their actionability (3). These systems may also vary in the types of data reported and frequency of reporting. In addition to discrepancies in real-time reporting of COVID-19 data, there may be differences in reporting of epidemiological risk factors and healthcare metrics. Accurate, comprehensive, and interoperable national data reporting systems are crucial for online databases that aggregate international data such as the WHO COVID-19 Dashboard and the COVID-19 Dashboard by Johns Hopkins University (4,5).

At the beginning of the COVID-19 pandemic, multiple African countries rapidly increased diagnostic and surveillance capacity (6). Additionally, most African countries quickly enacted nonpharmaceutical interventions to mitigate transmission, which came at a cost for healthcare systems and recovery (7). Given differences among African countries and varying experiences with COVID-19, there have been diverse practices in data reporting. The Africa Centers for Disease Control and Prevention (CDC) uses an online dashboard to aggregate COVID-19 cases, deaths, and recoveries from official regional collaborating centers and member states (8). Similarly, the Africa CDC has an online dashboard for national vaccination rates (9). However, countries also have their own national reporting systems which can provide more detailed data for researchers, clinicians, decision-makers, and the public. Such national reporting systems may be less accessible and vary in their frequency, content, and formatting, so it is important to compare them with each other.

Many initially wondered why there was a lower than expected morbidity and mortality due to COVID-19 in Africa, and whether lower reporting could be contributing (10). Comprehensive and reliable data reporting are key to understanding this observation as well as others. For example, patient cohorts from North America, Europe, and Asia primarily shaped the initial international understanding of epidemiological risk factors for COVID-19, and further data are needed to assess risk factors specific to populations in Africa (11). Moreover, measurements of health metrics such as cases, demographics, co-morbidities, healthcare capacity, and hospital-acquired infections are necessary to address health disparities and inequities. Limited reporting of such data can translate into a lack of meaningful representation in international models and decision-making. As African nations face future emerging infectious diseases and look to strengthen national and continental public health institutions (12), insights from national data reporting during the COVID-19 pandemic will be critical to consider.

Therefore, we aimed to compare digital reporting of COVID-19 cases, risk factors, and healthcare capacity between African countries in order to identify lessons for future pandemic response and preparedness.

## Methods

We searched for national COVID-19 reporting systems for 54 African countries during 2020 by reviewing governmental websites and social media postings belonging to national ministries of health and public health, as well as the source materials for aggregated COVID-19 dashboards. We included reporting systems that were created and/or endorsed by national governments. Reporting systems were divided by type of report, which included social media and website postings, press releases, situation reports, and online dashboards. Situation reports were defined as daily to monthly published electronic documents that had consistent formatting, figures, and data beyond total cases/deaths/recoveries. Online dashboards included visuals and additional data but were updated more frequently (often daily or in real-time). Press releases were text-based summaries that were posted infrequently. Social media activity included intermittent postings on Twitter and Facebook. Some social media accounts also included electronic press releases or situation reports. Websites were defined as internet sites that had minimal COVID-19 data, such as only total cases/deaths/recoveries. For the countries that had situation reports and/or online dashboards, we further compared whether there was reporting of different types of case data (total cases, deaths, recoveries, number tested, and healthcare workers infected), epidemiological data of cases (age, sex, and co-morbidities), and healthcare capacity data (number hospitalized, bed availability, and ventilator or oxygen availability). We also compared whether subnational data were reported and whether there was reporting of testing practices (polymerase chain reaction, serology, or rapid antigen testing). We also determined whether countries had a specific COVID-19 support fund according to the International Monetary Fund (IMF) (13). Lastly, we examined regional reporting practices from the five United Nations geoscheme sub-regions (Central, North, East, West, and Southern Africa). We further analyzed national reporting in Cameroon, Egypt, Kenya, Senegal, and South Africa as examples from these sub-regions.

## Results

All 54 African countries had diagnosed cases of COVID-19 according to aggregated dashboards (4,5,8). We were able to identify official national COVID-19 reporting systems in 53 of 54 countries (98.1%); Tanzania was the only country without a reporting system. Of the countries with reporting systems in 2020, 36/53 (67.9%) had recurrent situation reports and/or updated online dashboards. Of those with online dashboards or situation reports, 30/36 (83.3%) had subnational data, 20/36 (55.5%) included total number of tests performed, 19/36 (52.8%) included patient sex, 14/36 (38.9%) included patient age, and 5/36 (13.9%) reported patient co-morbidities. Social media was used by 37/54 (68.5%) of African countries to report official COVID-19 statistics. An extrabudgetary COVID-19 fund was identified in 27/54 (50%) of African countries. Comparing sub-regions, Southern Africa had the highest percentage of countries with situation reports and/or online dashboards, 4/5 (80%), followed by West Africa, 12/16 (75%), Central Africa, 6/9 (66.7%), East Africa, 11/18 (61.1%), and North Africa, 3/6 (50%).

A variety of reporting systems were found among African countries. Reporting systems varied in the types of data reported and the frequency of reporting. Most COVID-19 data reporting systems included total cases, deaths, and recoveries. Reporting systems increased in complexity and content from social media postings, to occasional website postings and press releases, to routine situation reports with graphs and figures, to online dashboards updated daily or in real-time. COVID-19 data reporting systems among African countries are shown in Figure 1, and the accompanying national sources are available in Table 1. Routine situation reports and online dashboards had the greatest breadth and frequency of data. Overall, there was great diversity in data reporting systems, with no clear standards for the content, frequency, or format of reporting. We have further examined and compared reporting practices by a few selected countries and their respective sub-regions below.

**Table 1.**
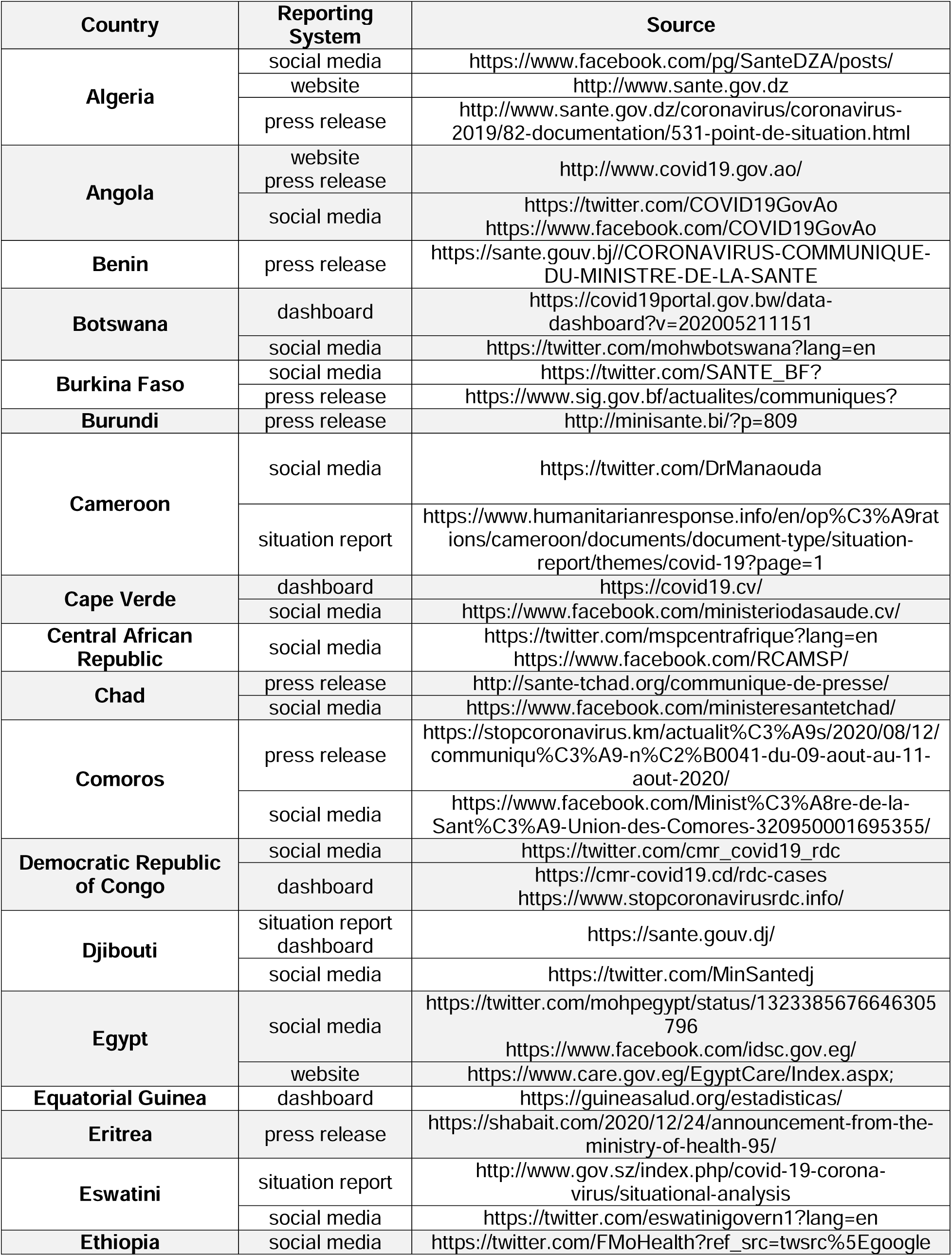
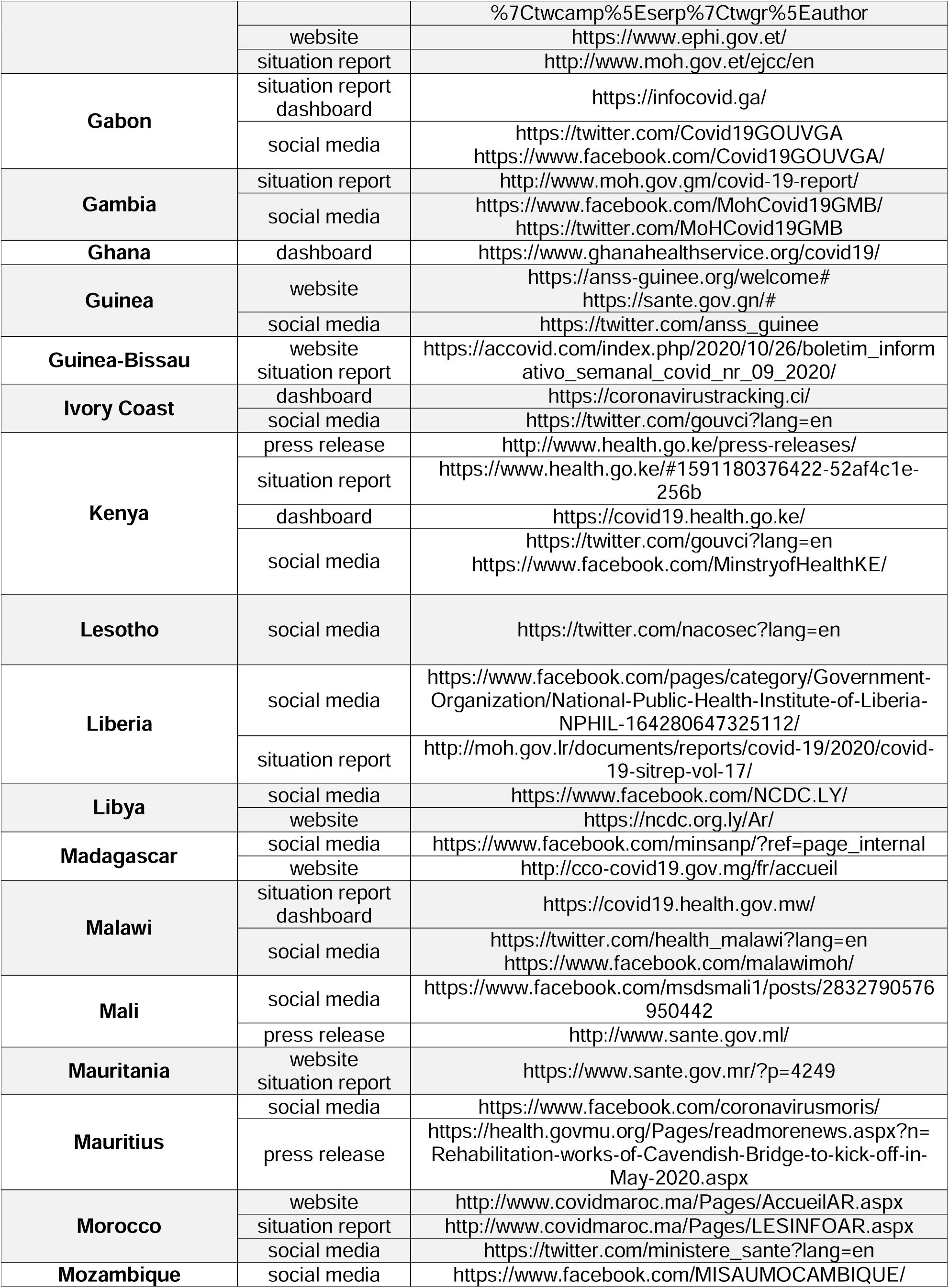
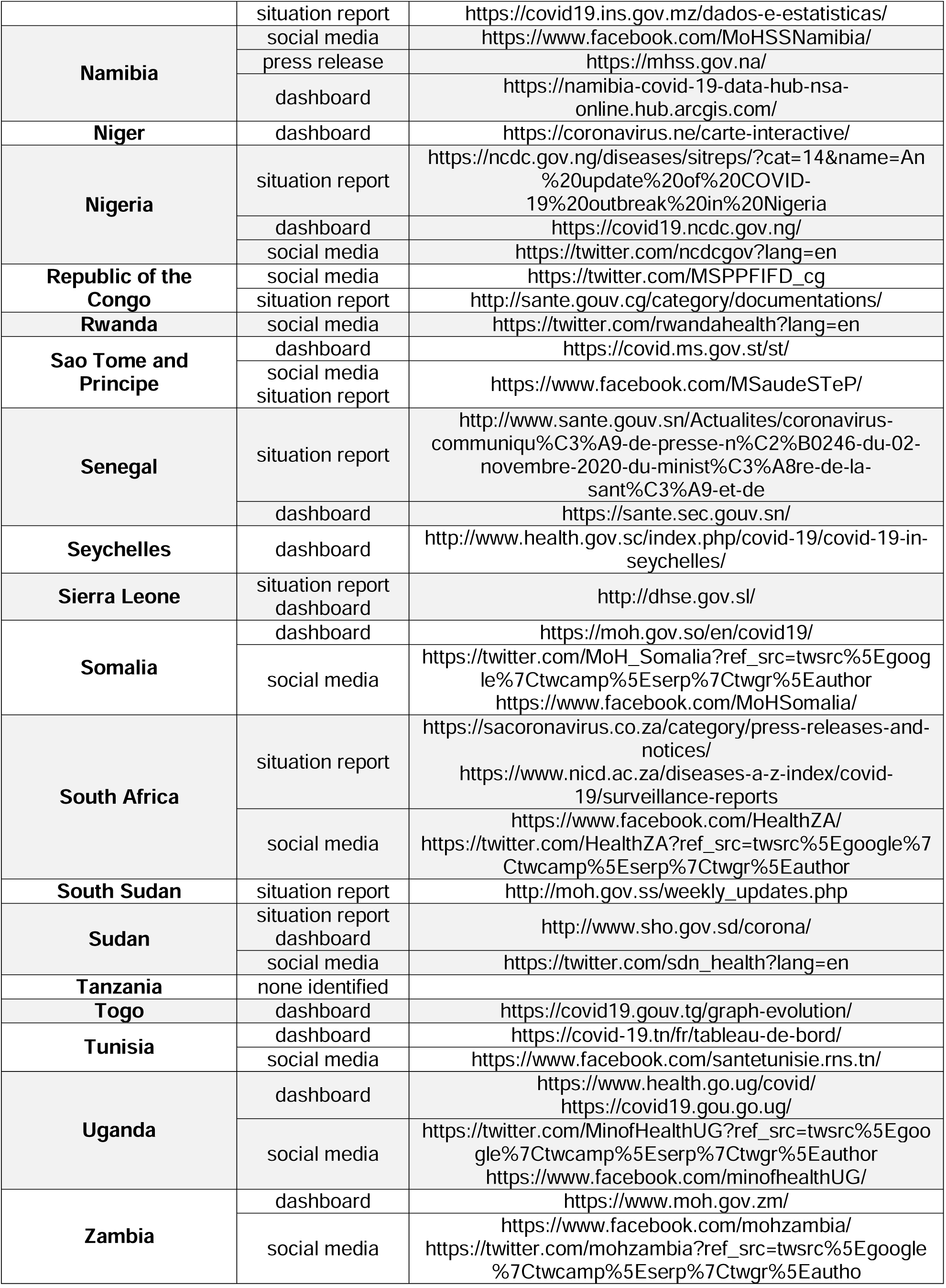
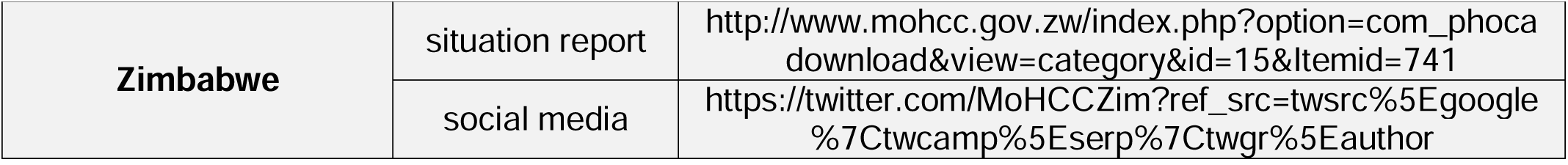
National sources for COVID-19 Data in Africa

**Figure 1.**
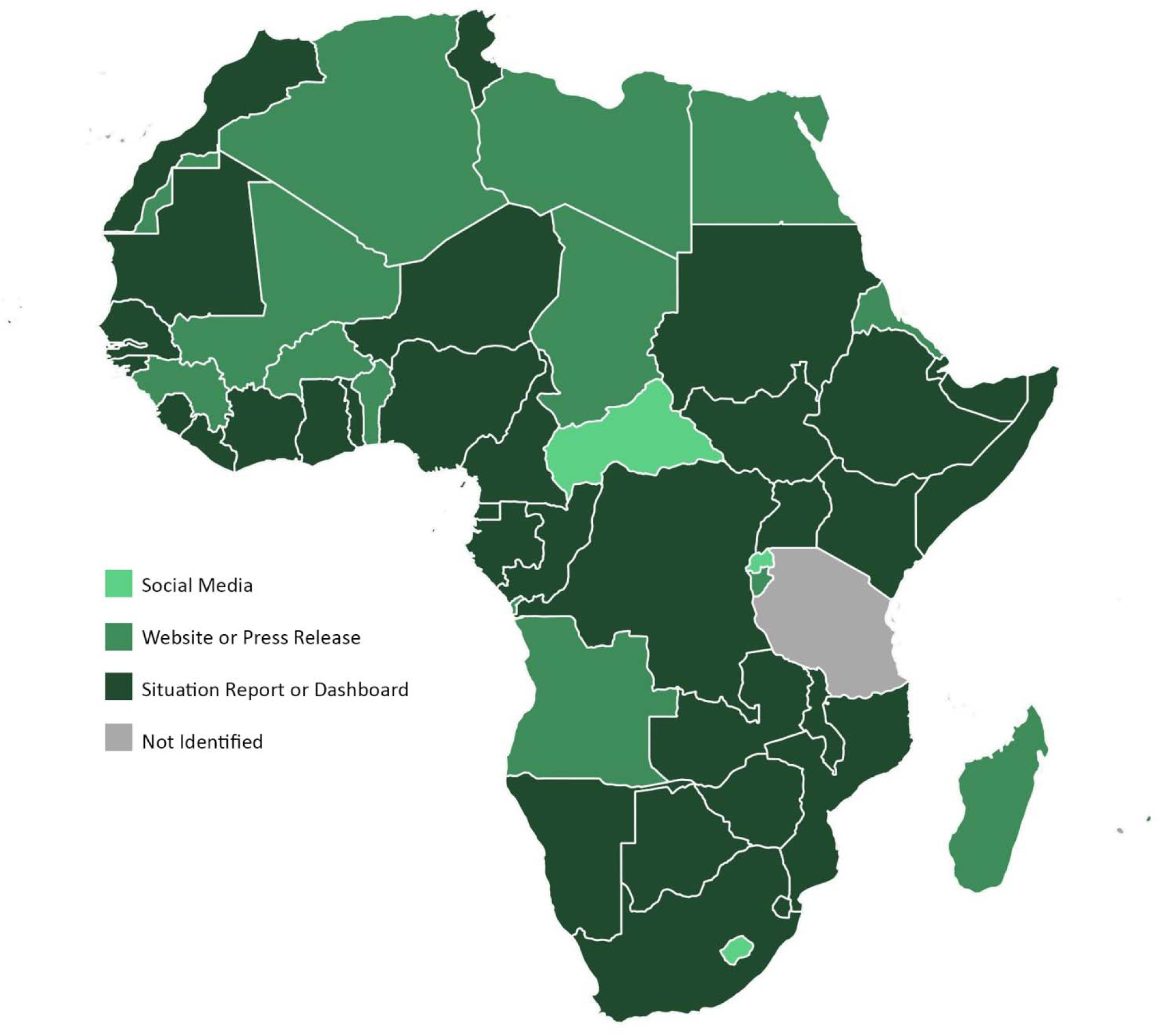
National COVID-19 data reporting systems in Africa African countries are depicted with identified type of national COVID-19 data reporting system. If a country used multiple reporting systems, the system with higher data quality is shown.

### Cameroon

On February 7^th^ 2020, the Ministry of Public Health in Cameroon released the first national situation report on COVID-19. Approximately 1 month later, the first case of COVID-19 was officially reported in Cameroon (14). As the pandemic spread throughout Cameroon, situation reports were published approximately weekly. The content of the reports evolved as the global situation grew and more clinical data became available. The first situation report containing healthcare capacity data was released on May 25^th^, while the first report with co-morbidity data was published on August 31^st^. Compared to other countries with COVID-19 reporting systems, Cameroon was one of the few countries that routinely reported metrics of healthcare capacity in their situation reports, including data on bed availability and oxygen concentrators. Cameroon was also one of the few countries to report cases among healthcare workers and diagnostic testing capacity. Situation reports from Cameroon were less publicly accessible, and reports had to be obtained from the Cameroon Coordination Center for Public Health Emergency and an online humanitarian response repository supported by the United Nations (15). Five other countries in Central Africa, including the Democratic Republic of the Congo, Equatorial Guinea, Gabon, Republic of the Congo, and Sao Tome and Principe, had online dashboards and/or situation reports.

### Egypt

The first confirmed case of COVID-19 in Africa was announced in Egypt on February 14^th^, 2020 (16). A month later, 33 passengers and 12 staff members on a Nile cruise ship were infected with SARS-CoV-2, prompting the Egyptian government to increase preventive measures (17). Early models comparing exported cases and the total number of reported cases by the Ministry of Health in Egypt predicted that the burden of COVID-19 was underreported (18). The Egyptian Ministry of Health reported total cases, mortalities, and recoveries on a website in Arabic. Data about demographics and co-morbidities were not available. The Ministry of Health also posted statistics on Twitter and Facebook. Only 3 of 6 North African countries: Tunisia, Morocco, and Sudan, were found to have COVID-19 situation reports or online dashboards, which provided more detailed data.

### Kenya

Kenya also utilized frequent situation reports and an online dashboard to report COVID-19 data. Situation reports were published almost daily during June and July of 2020. Unlike most other countries, Kenya also reported data on the total number of tests performed and the type of testing, which is important for understanding surveillance and diagnostic capacity. Additionally, Kenya reported demographics and co-morbidities of cases, as well as the distribution of cases by counties and sub-counties. Kenya also reported hospitalizations, Intensive Care Unit (ICU) admissions, and patients on home-based isolation. Early during the pandemic in Kenya, the distribution and availability of ICU beds and ventilators were a main concern (19). By mid-2020 Kenya had 537 ICU beds and 256 ventilators, with the majority of ICU beds found within Kenya’s major metropolises of Nairobi and Mombasa (20). The majority of countries in East Africa were found to have high-quality reporting systems. No public national reporting system could be identified in Tanzania, where controversies of governmental misinformation about COVID-19 arose early during the pandemic (21).

### Senegal

Senegal has been recognized for its overall response to the pandemic and performing well on a COVID-19 global response index (22). The Ministry of Health and Social Action in Senegal reported COVID-19 case data using both situation reports as well as an online dashboard that is updated in real-time. Compared to other sub-regions, West Africa had a high percentage of countries with high-quality COVID-19 reporting. One possible reason for this is the experience many of these countries gained in data reporting during the 2013-2016 Ebola virus disease epidemic in West Africa. Timely diagnostics, reporting of cases, and contact tracing were critical for both the Ebola virus disease epidemic and the COVID-19 pandemic (23).

### South Africa

South Africa has identified the most cases of COVID-19 on the African continent, with over 2,860,000 cases identified at the time of this report. The South African COVID-19 Online Resource & News Portal from the Department of Health has frequent updates on COVID-19 statistics. Additionally, the National Institute for Communicable Diseases provides daily hospital surveillance reports, which include more detailed clinical information. Hospitalizations, demographics, and co-morbidities of COVID-19 cases were all reported in South Africa. One of the first large cohort studies of co-morbidities in COVID-19 patients in Africa was reported from South Africa (24). Given that there have been few studies of co-morbidities in COVID-19 patients in Africa and national data reporting systems were found to rarely include co-morbidities, more research and data reporting are needed to understand epidemiological risk factors for COVID-19 in other regions of Africa. Additional countries in Southern Africa that had online dashboards and/or situation reports included Botswana, Eswatini, and Namibia.

## Conclusions

Overall, the diversity and differences among COVID-19 reporting systems in African countries reveal international lessons for data reporting during pandemics. Many countries adopted sophisticated, routine situation reports or online dashboards that could be updated in real-time. While most countries included counts for cases, deaths, and recoveries, a few countries reported additional data such as demographics, co-morbidities, testing, and healthcare capacity. These additional metrics can be useful for researchers, decision-makers, clinicians, and the general public to understand the current status of the pandemic. Accurate and accessible data on cases, epidemiology, and healthcare capacity will be important inputs for novel COVID-19 modeling tools in Africa (25). These data will also be critical for conducting vaccination campaigns and assessing populations at risk for COVID-19 sequelae.

One limitation of our study was that it focused on publicly available reporting systems during 2020, and we may have not have found official reporting systems that were less accessible or did not fit our classification paradigm. National reporting systems are essential to keeping the public well-informed and trusting of their government and therefore must be accessible. Lack of government trust has been identified as a reason for widespread vaccine hesitancy in African countries (26). Clinicians, researchers, and the general public are still experiencing the consequences of the Tanzanian government withholding COVID-19 data reporting early on in the pandemic (27).

National data reporting systems are important for transmitting relevant and reliable data during pandemics. While national data reporting may vary by context, standardizing data reporting systems could allow for greater comparison and dispersion of data. Certain groups that aggregate data for decision-making, such as the Africa CDC or WHO, could establish a standard for the minimum public national data required during a pandemic, which could be dynamic and adaptive as key questions emerge. Other countries could learn how to improve their own data reporting by examining the diversity of reporting systems in African countries. As we face future pandemics as an international community, developing and improving data reporting systems will be critical for unified pandemic preparedness and response.

## Data Availability

The authors confirm that the data supporting the findings of this study are available within the article and its supplementary materials. Additional data are available upon request from the corresponding author.

